# Introductions and early spread of SARS-CoV-2 in the New York City area

**DOI:** 10.1101/2020.04.08.20056929

**Authors:** Ana S. Gonzalez-Reiche, Matthew M. Hernandez, Mitchell Sullivan, Brianne Ciferri, Hala Alshammary, Ajay Obla, Shelcie Fabre, Giulio Kleiner, Jose Polanco, Zenab Khan, Bremy Alburquerque, Adriana van de Guchte, Jayeeta Dutta, Nancy Francoeur, Betsaida Salom Melo, Irina Oussenko, Gintaras Deikus, Juan Soto, Shwetha Hara Sridhar, Ying-Chih Wang, Kathryn Twyman, Andrew Kasarskis, Deena Rose Altman, Melissa Smith, Robert Sebra, Judith Aberg, Florian Krammer, Adolfo García-Sastre, Marta Luksza, Gopi Patel, Alberto Paniz-Mondolfi, Melissa Gitman, Emilia Mia Sordillo, Viviana Simon, Harm van Bakel

**Author notes:** Equally contributing authors (Ana S. Gonzalez-Reiche, Matthew M. Hernandez). Co-senior authors (Emilia Mia Sordillo, Viviana Simon, Harm van Bakel).

## Abstract

New York City (NYC) has emerged as one of the epicenters of the current SARS-CoV2 pandemic. To identify the early events underlying the rapid spread of the virus in the NYC metropolitan area, we sequenced the virus causing COVID19 in patients seeking care at the Mount Sinai Health System. Phylogenetic analysis of 84 distinct SARS-CoV2 genomes indicates multiple, independent but isolated introductions mainly from Europe and other parts of the United States. Moreover, we find evidence for community transmission of SARS-CoV2 as suggested by clusters of related viruses found in patients living in different neighborhoods of the city.

## INTRODUCTION

Severe acute respiratory syndrome coronavirus 2 (SARS-CoV-2; previously known as 2019-nCoV) is an emerging viral pathogen that was first reported to cause severe respiratory infections in Wuhan, China in late December 2019. Over the past months it has rapidly spread across the globe and the World Health Organization (WHO) declared a pandemic on March 11, 2020. Targeted screening of suspected coronavirus disease 2019 (COVID-19) cases, as well as a series of successive nation-wide travel restrictions were put in place to curtail SARS-CoV-2 introductions into the USA from outbreak hotspots in China (January 31, 2020), Iran (February 29, 2020), mainland European countries (March 11, 2020), and the UK and Ireland (March 14, 2020) to the continental United States. Despite these measures, the first SARS-CoV-2 case in New York State was identified in New York City (NYC) on February 29, 2020. Following a large expansion of screening capacity in the first weeks of March the number of cases rapidly increased, and as of April 5, 2020, there were 122,031 confirmed SARS-CoV-2 cases in NY state, including 67,551 (51%) in NYC (NY State Department of Health (NYS DOH); https://coronavirus.health.ny.gov/). With more than 3,000 fatalities in the metropolitan area, NYC has become one of the major epicenters of SARS-CoV-2 infections in the USA.

The Pathogen Surveillance Program (PSP) at the Icahn School of Medicine at Mount Sinai is a multidisciplinary, institutional infrastructure which seeks to generate high resolution, near real-time genetic information on pathogens found to cause disease in the large and diverse patient population seeking care at the Mount Sinai Health System in NYC. Following biospecimen coding, nucleic acid extraction and qPCR quantification, cutting edge next-generation sequencing approaches based on Illumina and PacBio technology provide information on the pathogen’s genome. The process has been optimized for quick turnaround, optimized data assembly and, last but not least, integration with de-identified epidemiological information.

We took advantage of the PSP to investigate the origins of SARS-CoV-2 strains circulating in NYC and to dissect its spread in this metropolitan area with a high-density population. Here we present the genomic diversity of 90 SARS-CoV-2 isolates obtained from 84 patients seeking care at the Mount Sinai Health System between February 29, 2020 and March 18, 2020. These genomes provide clear evidence for multiple, independent SARS-CoV-2 introductions into NYC during the first weeks of March. Based on genetic similarity and phylogenetic analysis of full-length viral genome sequences, most cases diagnosed during the 18 days after the first-reported case appear to be associated with untracked transmission and potential travel-related exposures. Cases with a history of travel exposure showed no evidence of onward transmission, supporting the efficacy of self-quarantine measures. Notably, the majority of introductions appear to have been sourced from Europe and the USA. We also identified two clusters totaling 21 closely related cases suggesting community spread. These observations are also supported by the city-wide distribution of these cases, which mapped to three of four represented NYC boroughs and five NY State Counties. Our data provide insight into the limited efficacy of travel restrictions for preventing spread of SARS-CoV2 infections in NYC, and point to the increased risk of community driven transmission within the larger metropolitan area.

## RESULTS

### Sequenced isolates represent SARS-CoV-2 cases from multiple locations across New York City area

Initial diagnostic testing for SARS-CoV2 in NYC was targeted and limited to individuals fitting a set of criteria outlined by CDC, and requiring pre-approval by the local Department of Health. Between March 9-14, screening capacity at the Mount Sinai Health System (MSHS) was greatly expanded, leading to a surge of newly diagnosed cases (**Figure 1**). Within one week, the number of daily positive SARS-CoV-2 tests exceeded the normal volume of positive tests for influenza virus by a factor of 5. We sequenced 90 SARS-CoV-2 genomes from multiple clinical isolates taken from 84 of the >800 cases identified up to March 18, yielding 72 complete and 18 partial (>95% coverage) genomes. These cases were drawn from 21 NYC neighborhoods across four boroughs (Manhattan, Bronx, Queens and Brooklyn), as well as two towns in Westchester County. Sequenced isolates were obtained directly from nasopharyngeal swab samples collected from 44 females (52.4%) and 40 (47.7%) males ranging in age between 20-44 years (16%), 45-64 years (33%), 65-79 years (27.4%), and >80 (16.7%) years. Of the 68 cases with available hospital visit information, 12 were discharged (17.6%), 53 were admitted (80%), and 3 were initially discharged and later admitted on a subsequent visit (2.4%).

**Figure 1.**
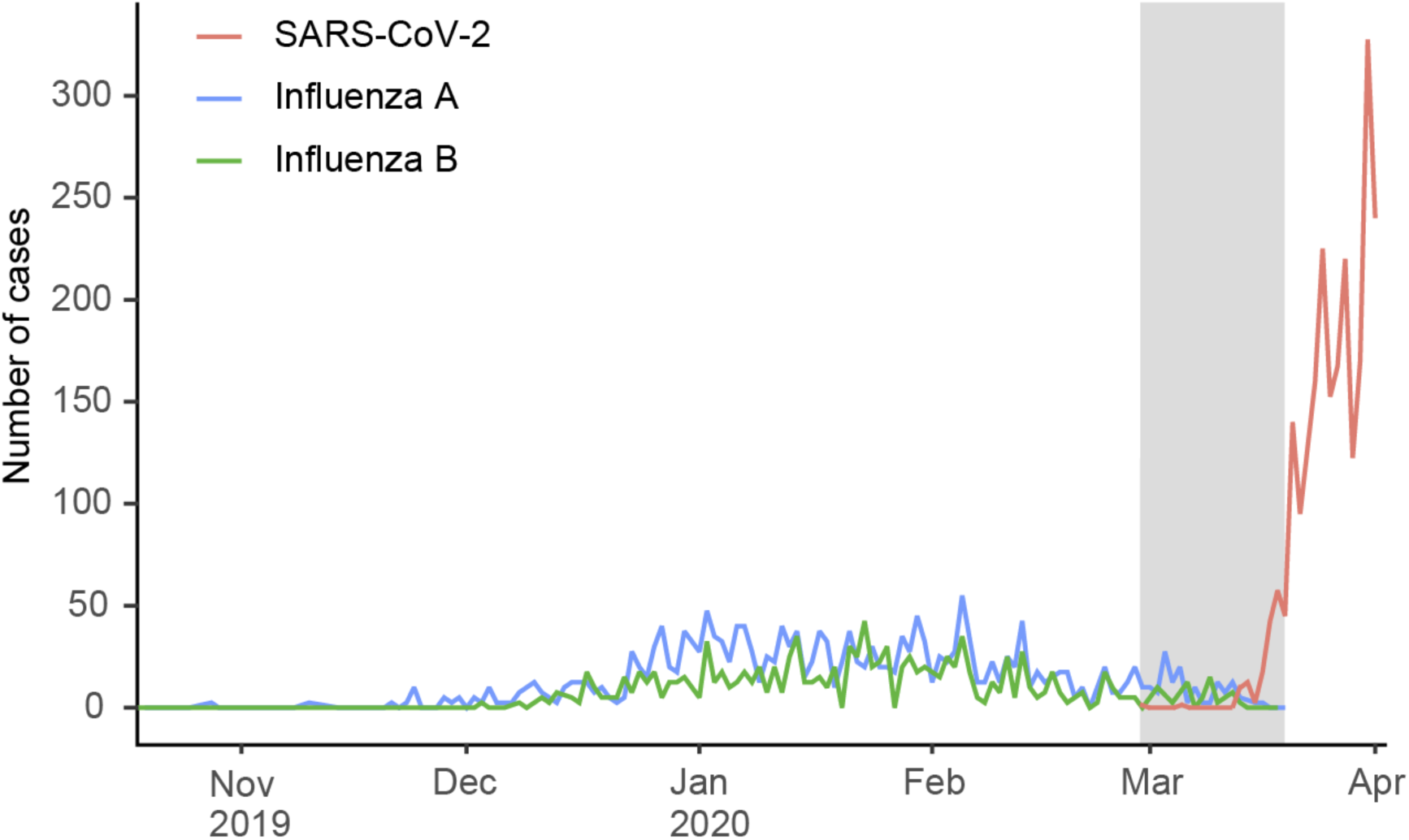
SARS-CoV-2 confirmed cases. Number of patients with positive molecular tests for SARS-CoV-2 up to March 31 2020, compared to the daily number of patients with influenza A and B tests in the 2019-2020 season. The shaded area indicates the period in which the samples sequenced in this study were obtained (February 29th to March 18). The number of positive tests per virus was not normalized for the number of tested samples.

### Phylogenetic analysis reveals multiple introductions to NYC from diverse origins

We performed phylogenetic analysis of the 84 unique patient isolates, together with 2,363 sequences deposited in GISAID up to April 1, 2020 (**Figure 2A**). NYC isolates were distributed throughout the phylogenetic tree; consistent with multiple independent introductions. We assigned each SARS-CoV2 isolate to main monophyletic clades based on amino acid and nucleotide substitutions and statistical support (see Methods). These mutations were used only for the purpose of classification, as the functional impact of many of these substitutions remains unknown. For each assigned clade, we identified different types of events according to the position of the NYC sequences and available epidemiological information (**Table 1**). The first isolate had documented exposure through travel to the Middle East (clade A3) and the second through travel to Europe (clade B). Neither of these showed evidence of onward transmission, so we therefore excluded these two cases from any inference made from the phylogenetic analyses. For the remaining isolates, the great majority (87%) cluster with clade A2a. This clade is largely composed of isolates obtained from patients with COVID19 in Europe (72%; **Figure 2B**), suggesting that introductions from Europe account for the majority of cases found in NYC in the first weeks of March 2020.

**Table 1.** Inferred SARS-CoV-2 virus transmission events related to New York City, March 2020

| Clade | Mutations | First NYC sequence | Cases (this study) | type of event | Inferred source | Inferred tMRCA (90% CI)* |
| --- | --- | --- | --- | --- | --- | --- |
| A1a | ORF3a:G251V and C14805T | 4-Mar | 5 | untracked transmission | Europe | Jan-28<br>(Jan-19 to Feb-17) |
| A2a | ORF1b:P314L | 11-Mar | 69 | untracked transmission | Europe / North America | Jan-25<br>(Jan-19 to Feb-02) |
|  |  |  | 4 | untracked transmission | Europe / North America | Feb-20<br>(Jan-08 to Feb-20) |
| A3 | ORF1a:V378I | 29-Feb | 1 | travel exposure | Middle East (epi link) | test date (Feb-29) |
| B | ORF8:L84S | 4-Mar | 1 | travel exposure | Europe (epi link) | test date (Mar-4) |
|  |  |  | 1 | putative introduction to NY | Europe | Feb-20<br>(Jan-29 to Feb-26) |
|  |  |  | 1 | putative introduction to NY | North America / Domestic | Mar-12<br>(Mar-06 to Mar-16) |
| B1 | C18060T | 17-Mar | 1 | putative introduction to NY | Domestic (WA) | Mar-03<br>(Feb-26 to Mar-12) |
| B4 | N:S202N | 14-Mar | 1 | putative introduction to NY | Asia/Oceania | Mar-03<br>(Feb-25 to Mar-12) |

\*Inferred time of the most recent common ancestor (tMRCA) at the base of the node containing NY-origin with >70% bootstrap support value. Travel exposures and putative introductions are shown for the cases described in this study.

**Figure 2.**
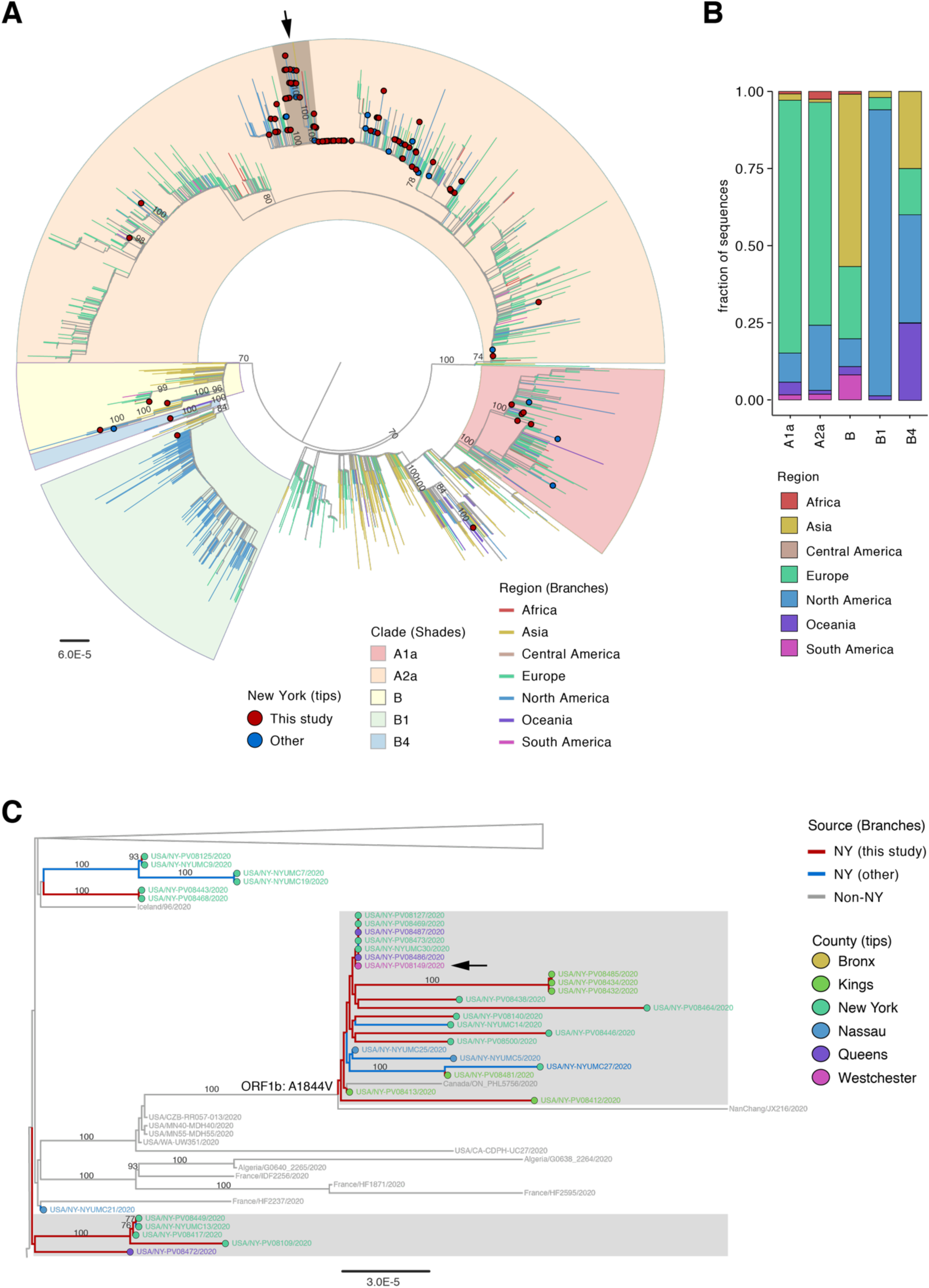
Phylogenetic relationships of SARS-CoV-2 from NY and other global strains. **A)** Maximum Likelihood phylogeny of 2,363 global SARS-CoV-2 sequences available in GISAID EpiCoV database as of April 1, 2020 and 84 NY sequences from this study. Branches are colored according to the region of origin. Tip circles indicate the position of NY sequences in red (this study) and blue (other NY sequences). Clades that contain NY sequences are highlighted; the local transmission clusters are indicated by the arrows. **B)** Stacked barplot showing the fraction of sequences per region by clade. **C)** Local transmission clusters on the ML tree showing the source of cases by county. Bootstrap support values >70% are shown only for main clades and clusters containing NY cases, sister clusters are collapsed for easier visualization. The mutation identified specific to the community transmission cluster is indicated. Scale bar at the bottom of panels A and C indicates number of nucleotide substitutions per site.

Most of the NYC isolates within clade A2a were interspersed and we did not observe grouping by country or geographical region (**Figure S1**). Despite their diverse origins, many sequences in this clade are highly similar or identical, which makes it impossible to resolve direct relationships or directionality between them. By placing the ML phylogeny on a time scale, we inferred a period of untracked global transmission between late January to mid-February, consistent with epidemiological observations of the developing pandemic. The earliest sequences at the base of clade A2a include isolates from Italy, Finland, Spain, France, the UK, and other European countries from late February, in addition to a few North American isolates (Canada and US) from the first week of March 2020.

Similar to isolates in clade A2a, isolates in our study positioned in clade A1a (6%) were interspersed among isolates from multiple regions with unknown directionality (**Figure 2B**). This clade is also largely composed of European-origin isolates (82%) where many of the early isolates at the base of the clade are from the UK.

For the rest of the clades (B, B1, and B4), we identified four putative SARS-CoV-2 virus introductions to NYC as early as February 20 (90%CI: January 29 to February 26) (**Table 1**). Notably, two of these introductions were inferred to be of domestic origin based on their close relationship with US isolates, including those from the main community transmission in Washington state (Clade B1) (Bedford et al. 2020). The introduction of this clade to the East Coast was recently reported (Fauver et al. 2020). Although more than half of the sequences in clade B were of Asian origin (**Figure 2B**) the closest relatives to the NY isolates were of European and North American origin. The isolate that belongs to clade B4 is positioned in a cluster with two US sequences from WA state, with an inferred date of introduction to NY in early March (**Table 1**) and a prior period of untracked transmission in unknown location(s) since January 21 2020 (90% CI: January 18 to January 23). Prior to this period, the closest viral isolates basal to this cluster are from Australia and China (**Figure S1**).

The sequenced isolates and assigned clades were spatially distributed throughout all NYC boroughs and 21 neighborhoods (**Figure 3**). Isolates sequenced at other NYC hospitals collected within a similar time window (March 3 to 17, 2020) belong to clades A1a, A2a, and B and were mainly collected in Manhattan (23 out of 31) and Nassau County (5 out of 31) based on the accompanying metadata available in GISAID (e.g., submission from NYU Langone Health, EPI_ISL accession no. 414639, 416830-32, 418190-205, 418254, and 418968-980 by Arguero-Rosenfeld, Black, Chen, et al., 2020).

**Figure 3.**
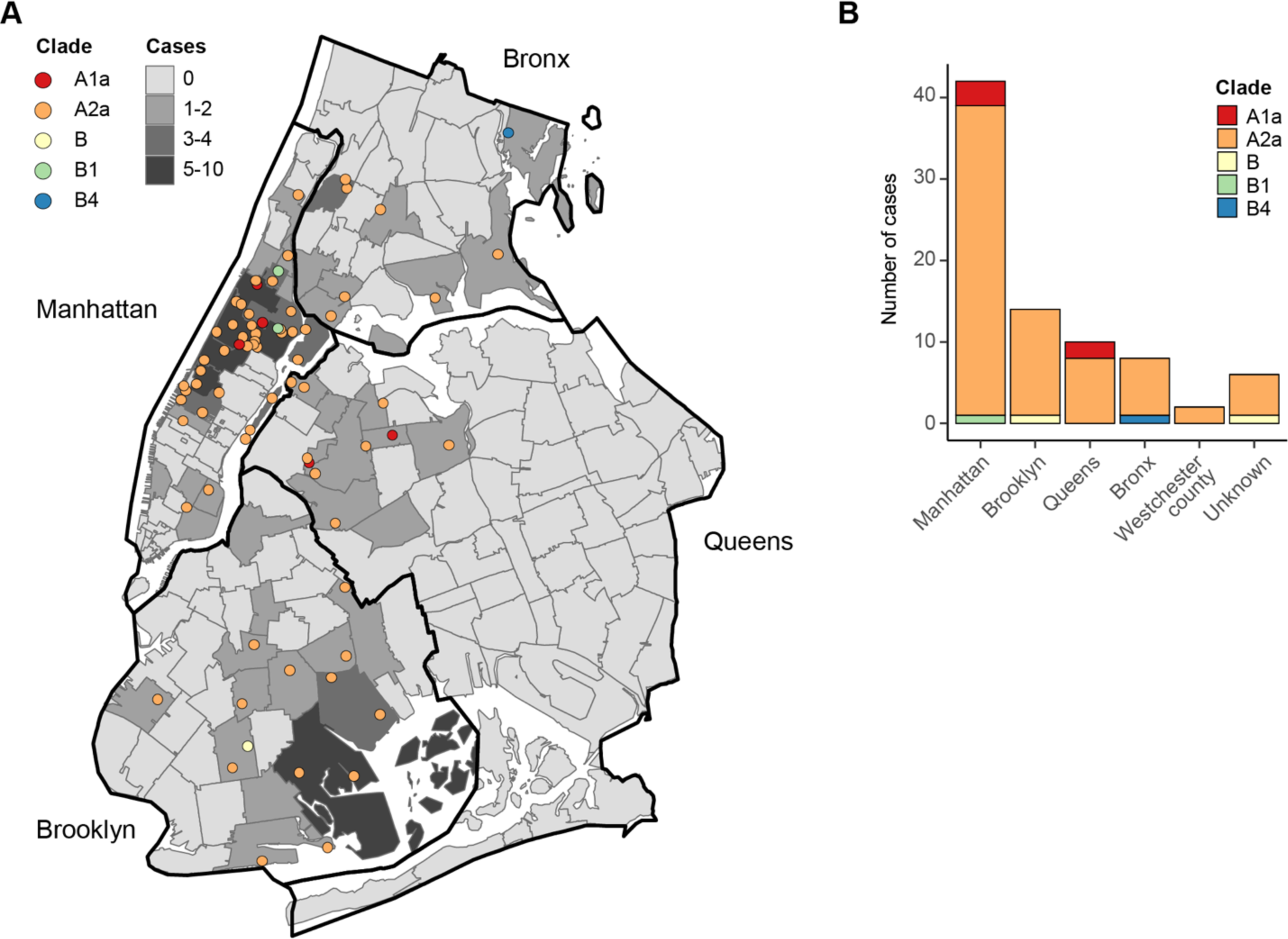
Distribution of the geographic location of the patients with COVID-19 from which viral isolates were sequenced. **A)** Distribution of 74 sequenced cases with available zip code information across NYC boroughs and neighborhoods. Each point represents one case and is colored according to the phylogenetic clade of the SARS-CoV-2 genome (see also Figure 2A). Neighborhoods are shaded according to the number of cases that were sampled. **B)** Breakdown of sequenced cases according to phylogenetic clades across NYC boroughs and Westchester county. Cases without available zip code information are indicated as ‘Unknown’.

Taken together, our results show that the NYC SARS-CoV-2 epidemic has been mainly sourced from untracked transmission between the US and Europe, with limited evidence of direct introductions from China where the virus originated.

### Community spread throughout NYC is mainly linked to European clades

Despite the relatively small number of SARS-CoV-2 sequences available to date, we identified two monophyletic clusters positioned within clade A2a that contained almost exclusively isolates from NY (**Figure 2B**). One cluster included 23% (17) of the isolates contained in clade A2a and 20% of the total isolates sequenced. Based on zip code information, the cases from this cluster were distributed across five counties, including one sample from New Rochelle, Westchester County, where the first cluster of community-acquired infections in NY state was found on March 5, 2020. This cluster was characterized by the amino acid substitution A1844V in the ORF1b gene. Basal to these clusters are isolates from the states of MN, WA, and CA. The time of the most recent common ancestor between other US and NY isolates suggests that viruses from the A2a clade could have been introduced to NY as early as February 29 (lower 90% CI limit), through a domestic route (**Figure S1**).

The second cluster was a smaller group that contained four isolates from Manhattan/NYC and one isolate from Queens/NYC. Zip code information was available for three of the Manhattan cases, which were mapped to three different neighborhoods further supporting community spread. Although most NYC cases are intermixed within this largely European clade, these results suggest that domestic introductions may have also been the source of early community spread within NYC.

## DISCUSSION

SARS-CoV-2 is the cause of one of the largest non-influenza pandemics of this century. Sequencing of SARS-CoV-2 isolates from around 10% of the patients diagnosed with COVID19 at the largest healthcare system of NYC during the first weeks of March provides unexpected insights into the origin and diversity of this new viral pathogen. We find clear evidence for multiple independent introductions into the larger metropolitan area from different origins in the world as well as the USA. These early introductions failed to seed larger clusters suggesting that early quarantine, hospitalization and contact tracing in the early days limited further spread. With increased testing, we observed the emergence of community acquired infections with the majority of the community cases caused by viral isolates from clades that are of European origin.

A limitation of our analysis is the relatively small number of isolates from cases identified in the first week of March 2020 which means that our model relies on inferences based on sequences deposited in the GISAID database. Since sequencing efforts vary by country, the fraction of sequences available by region/country is not necessarily representative of the number of cases reported for each of these regions. Thus, some of these inferences may change as more complete and representative SARS-CoV-2 sequences become available.

Taken together, we provide a first analysis of the SARS-CoV-2 viral genotypes collected from patients seeking medical care. We find that New York City, as an international hub, provides not only a snapshot of the diversity of disease-causing SARS-CoV-2 at the global level but also informs on the dynamics of the pandemic at the local level. Future studies are needed to define viral phenotypes and explore the impact of the public health measures such as closing of educational facilities (March 18, 2020), closing of non-essential services (March 20, 2020) and stay at home orders (March 22, 2020) on community transmission.

## METHODS

### Ethics statement

This study was reviewed and approved by the Institutional Review Board of the Icahn School of Medicine at Mount Sinai.

### Collection of nasopharyngeal swab samples that tested positive for SARS-CoV-2

Individuals infected with SARS-CoV-2 in NYC were initially identified through targeted screening of individuals with fever or signs/symptoms of respiratory illness, and either a history of travel to geographic outbreak hotspots (China, Japan, Italy, South Korea, Iran), or close contact with a confirmed case of COVID-19 within the past 14 days. Screening was also performed for cases with unknown exposure that had signs of severe respiratory illness requiring hospitalization. Authorization of the emergency use of an in vitro diagnostic tests for detection of SARS-CoV-2 virus and/or diagnosis of COVID-19 infection under section 564(b)(1) of the Act, 21 U.S.C. 360bbb-3(b)(1) enabled additional testing capacity to be rapidly deployed in hospital-based and private clinical laboratories during the first weeks of March. Tests for SARS-CoV-2 were performed in the MSHS Clinical Microbiology Laboratory, at Labcorp, the NY City Department of Health and Mental Hygiene (NYC DOHMH), or at the NY State Department of Health (NYS DOH). Labcorp or in-house testing used the Roche Cobas 6800 System or the Roche LightCycler 480 II.

For tests performed using the Roche Cobas 6800 System; the assay targets ORF1, a region that is unique to SARS-CoV-2. Additionally, a conserved region in the E-gene was chosen for pan-Sarbecovirus detection including SARS-CoV-2. Detection of the E-gene without detection of ORF1 is interpreted at a Presumptive Positive result.

For tests performed using the Roche LightCycler 480 II; nucleic acid was extracted on the EZ-1 Advanced XL or QIAcube Connect (QIAGEN) and Real-Time RT-PCR was performed using LightCycler 480 II (Roche) and 2019-Novel Coronavirus (2019-nCoV) primers and probes (IDT). The assay targets three regions of the virus nucleocapsid (N) gene. Amplification of all three targets is required to call a sample positive. The limit of detection of the assay is 1 genome copy/μL.

### Total RNA extraction

Total RNA was extracted from clinical specimens (e.g., nasopharyngeal swabs) using one of two methods. For manual extractions, 280 μL of viral transport medium from nasopharyngeal swabs was lysed with AVL buffer and viral RNA was extracted using the QIAamp Viral RNA Minikit (QIAGEN, cat. 52904), per the manufacturer’s instruction. For high-throughput specimen processing we used the KingFisher Flex Purification System (ThermoFisher, cat. 5400610) with the MagMax mirVana Total RNA Isolation Kit (ThermoFisher, cat. A27828) to extract total RNA from 250 μL of viral transport medium, as per the manufacturer’s protocol.

### Confirmatory testing and viral load determination by qRT-PCR

To detect SARS-CoV-2 RNA from clinical specimens, a modified version of the CDC 2019-nCoV real-time RT-PCR Panel was used (CDC/NCIRD/DVD). Primers and probes were commercially developed and comprised the 2019-nCoV Kit (Integrated DNA Technologies, cat. 10006606). Briefly, 2019-nCoV primer and probe sets consisted of two 2019-nCoV-specific sets (N1, N2) and one SARS-like universal set (N3) (**Table S1**). A fourth set was designed to detect host cellular RNaseP. Reactions were run using the QuantiFast Pathogen RT-PCR +IC Kit (QIAGEN, cat. 211454). Prior to quantitating SARS-CoV-2 RNA in clinical specimens, amplification efficiencies and limit of detection were assessed using six dilutions of commercially available plasmid controls (2019-nCoV_N_Positive Control, IDT, cat. 10006625; Hs_RPP30 Positive Control, IDT, cat. 10006626). Assays were run using USA/WA-1/2020 SARS-CoV-2 RNA as a positive control (25,000 genome copies per reaction) and nuclease-free water as a non-template control in a 384-well format. All four RT-PCR reactions were performed for each sample in duplicate using the following cycling conditions on the Roche LightCycler 480 Instrument II (Roche Molecular Systems, 05015243001): 50°C for 20 min, 95°C for 1 sec, 95°C for 5 min, followed by 45 cycles of 95°C for 15 sec and 60°C for 45 sec, during which quantitation of products (FAM) occurred.

All four RT-PCR reactions amplified with efficiency >0.9999 (R2≥0.9413) and the coronavirus primer-probe sets did not cross-react with viral RNA from clinical specimens containing other coronavirus strains (e.g., OC43, NL63, HKU1) or influenza A. For a specimen result to be positive for SARS-CoV-2, all three coronavirus reactions (N1, N2, N3) were required to be positive (Ct < 38), regardless of the quantity of RNaseP detected. A result was deemed negative if all three reactions failed to detect the product (Ct ≥ 38) and there was sufficient input (e.g., RNaseP Ct < 35). A result was deemed inconclusive if one or two of the three failed to amplify (Ct ≥ 38). A result was deemed invalid if all three coronavirus reactions were negative and if there was evidence of insufficient input (e.g., RNaseP Ct ≥ 35). SARS-CoV-2 genome copy number in extracted RNA was quantitated by comparing the average coronavirus Ct across N1, N2, and N3 to that of the positive control.

### Whole-genome amplification and sequencing

Sample preparation for sequencing was done using whole-genome amplification with custom designed tiling primers (Quick et al. 2017) and the Artic Consortium protocol (https://artic.network/ncov-2019), with modifications. In brief, cDNA synthesis with random hexamers was performed with ProtoScript II (New England Biolabs, cat. E6560) from 7 μL of RNA according to manufacturer’s recommendations. The RT reaction was incubated for 30 mins at 48C, followed by enzyme inactivation at 85C for 5 mins. Targeted amplification was done from 5 μL of cDNA with the Q5 Hot Start High-Fidelity DNA polymerase (New England Biolabs, cat. M0493) (25 μL total reaction volume) with two sets of tiling primers generating ∼1.5 and ∼2 kb amplicons with ∼200 bp overlaps between each region. The PCR amplification parameters were: 1 min at 98C, 30-35 cycles of 15s at 98C and 5 min at 63C, and final extension for 10 min at 65C. Tiling primers were initially selected using Primal Scheme (Quick et al. 2017) and were further optimized in successive experiments to improve whole-genome coverage and assembly contiguity. The final list of optimized primers is provided in **Table S2**.

Amplicons were visualized on a 2% agarose gel and cleaned with 1.8X volume of Ampure XT beads. Amplicon SMRTbell libraries were prepared as recommended by the manufacturer (Pacific Biosciences). Briefly, the 1.5kb and 2.0kb amplicons were equimolar pooled, amounting to 500 nanograms of cDNA. The pooled amplicons were then used as input for SMRTbell preparation using the Express Template Preparation Kit 2.0. Each sample was first treated with a DNA Damage Repair mix to repair nicked DNA, followed by an End Repair and A-tailing reaction to repair blunt ends and polyadenylate each template. Next, overhang SMRTbell adapters were ligated onto the polyA track of each template and cleaned with 0.6X AMPure PB beads to remove small fragments and excess reagents (Pacific Biosciences). Amplicon libraries for Illumina sequencing were prepared using the Nextera XT DNA Sample Preparation kit (Illumina, cat. FC-131-1096), as recommended by the manufacturer.

For PacBio sequencing, completed SMRTbell libraries were annealed to sequencing primer v4 and bound to sequencing polymerase 3.0 before being sequenced using one 1M SMRTcell on the Sequel 1 system with a 20-hour movie. For Illumina, paired-end sequencing (2×150 nt) was performed on a MiSeq instrument.

### SARS-CoV-2 genome assembly

A custom reference-based analysis pipeline (https://github.com/mjsull/COVID_pipe) was used to reconstruct SARS-CoV-2 genomes. For genomes sequenced on the PacBio Sequel II platform, high-quality (>Q20) circular consensus sequence (CCS) reads were first generated from raw subreads, using the *SMRTLink* analysis suite, v8.0 with a minimum of 3 passes. Primer sequences were then trimmed from the ends of PacBio CCS long-reads or Illumina short-reads using cutadapt version 2.8 (Martin 2011). Trimmed reads were aligned to SARS-CoV-2 genome MN908947.3 using *minimap2* version 2.17-r941 (Li 2018), and consensus sequence was called using *Pilon* v1.23 (Walker et al. 2014), allowing for all variant types (single nucleotide variants, small insertions/deletions, large insertions/deletions or block substitution events, and local misassemblies). Regions with less than 10 fold-coverage were masked and unamplified regions at the end of the viral genome were also removed. Finally, consensus sequences were annotated using *prokka* v1.14.6 (Seemann 2014) and a custom SARS-CoV-2 reference annotation file.

### Phylogenetic analysis

Global background sequences were downloaded from GISAID EpiCoV database as of [April 1, 2020, accession numbers EPI_ISL_402119 to EPI_ISL_418989]. Only complete sequences with >75% of non-ambiguous sites and that contained complete date information (YYYY-MM-DD) were included. The alignment was done with *MAFTT* v7.455 (Katoh and Standley 2013) as implemented in the *NextStrain* tool (Hadfield et al. 2018). The alignment was manually curated to remove potential artifacts and trim 5’ and 3’ ends to further remove ambiguous regions. Duplicate sequences (i.e. identical sequences from the same location and collection date) were also removed. Maximum Likelihood phylogeny was inferred under the best-fit model of nucleotide substitution GTR+F+I+G4 in *IQTree* (Nguyen et al. 2015; Hoang et al. 2018). Tree topology was assessed with the fast bootstrapping function with 1000 replicates.

The ML tree was inspected in *TempEst* for outliers that deviated from a temporal root-to-tip divergence (Andrew Rambaut et al. 2016). The final inferred tree containing 2451 taxa, was time-scaled with *TreeTime* (Sagulenko, Puller, and Neher 2018) using a fixed clock rate of 8×10^−4^ (stdev = 4×10^−4^) under a skyline coalescent clock model, and rooted to minimize residuals on a root-to-tip and sample time regression (least-squares method). Marginal date estimates of ancestral states were inferred with 90% confidence intervals. Tree visualization and annotations were done in *FigTree* v1.4.4 (A. Rambaut 2014).

For both, introduction or transmission events were counted only when nodes had ≥70% bootstrap support value. Introduction events were defined as NY sequences that clustered with non-NY (US or other countries) sequences across different clades. Local transmission events were defined as NY-exclusive clusters with at least 5 or more sequences that were reproduced in the time-scaled inference.

Major clades that contained NY sequences were defined by amino acid and/or nucleotide substitutions with ≥70% bootstrap support value in the ML tree. For simplicity, clades were matched to the NextStrain nomenclature for SARS-CoV-2 as of April 4, 2020. (https://nextstrain.org/ncov).

### Spatial and statistical analysis

Spatial visualization was performed using zip-code data points based on patient addresses. The data was aggregated based on county, NYC borough, or NYC neighborhoods using publicly available zip-code definitions provided by the NYC Department of Health and Mental Hygiene. Maps were created in *ArcGIS Desktop* using the *ArcMap* v10.6 application available through the Environmental Systems Research Institute (ESRI). Baseline analysis across clades was completed using the R *tableone* package.

## Data Availability

SARS-CoV-2 genome sequencing data for all study isolates has been deposited in GISAID.

https://www.gisaid.org/

## Data availability

SARS-CoV-2 genome sequencing data for all study isolates has been deposited in GISAID with “NY-PV” strain names, accession numbers EPI_ISL_414476, EPI_ISL_415151, and EPI_ISL_421348 to EPI_ISL_421435.

## ACKNOWLEDGEMENTS

We are greatly indebted to our clinical colleagues for their dedication and courage in providing continued high-quality medical care under very difficult conditions.

We gratefully acknowledge the authors, originating and submitting laboratories of sequences from GISAID’s EpiFlu and EpiCoV (www.gisaid.org) that were used as background for our phylogenetic inferences. Special thanks to our colleagues at the Department of Pathology and Medicine at NYU School of Medicine (Maria Aguero-Rosenfeld et al) and at NYU Langone Health lab for submitting sequences from NYC to GISAID.

The Research reported in this paper was supported by the Office of Research Infrastructure of the National Institutes of Health (NIH) under award numbers S10OD018522 and S10OD026880 as well as institutional funds. Protocols established for this study were in part based on influenza virus sequencing protocols established with the support of CRIP (Center for Research on Influenza Pathogenesis), an NIH funded Center of Excellence for Influenza Research and Surveillance (CEIRS, contract number HHSN272201400008C). The content is solely the responsibility of the authors and does not necessarily represent the official views of the NIH.

## SUPPLEMENTARY MATERIALS

**Table S1.**
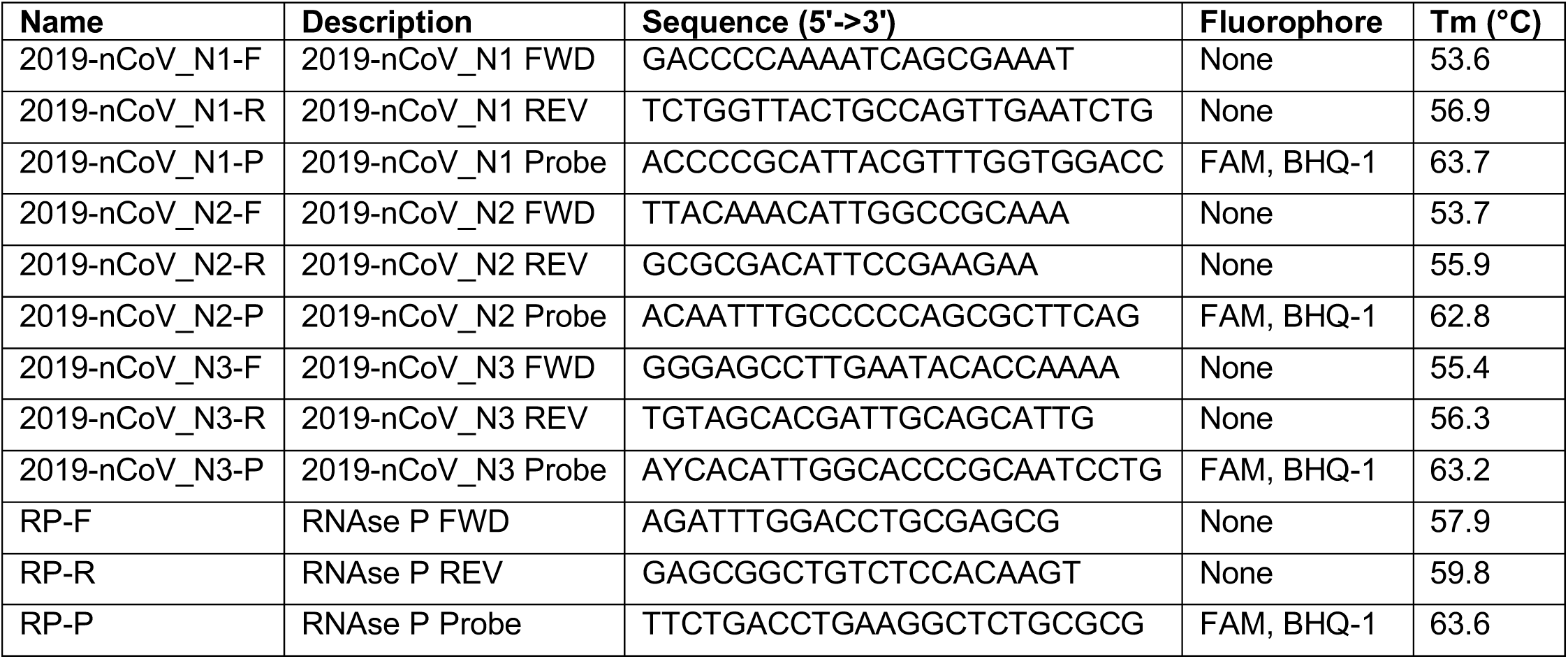
Primers for SARS-CoV-2 confirmation testing

**Table S2.**
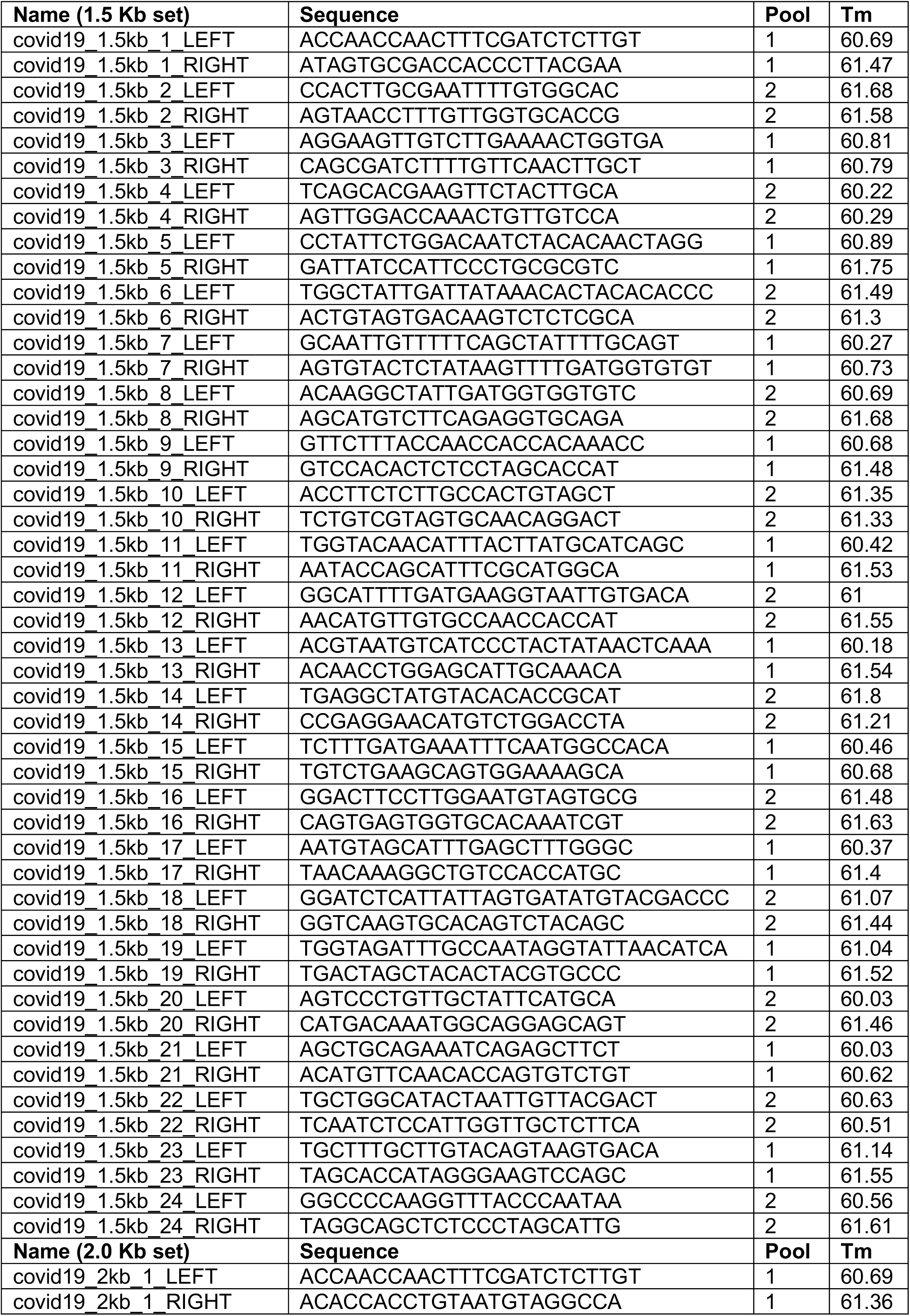
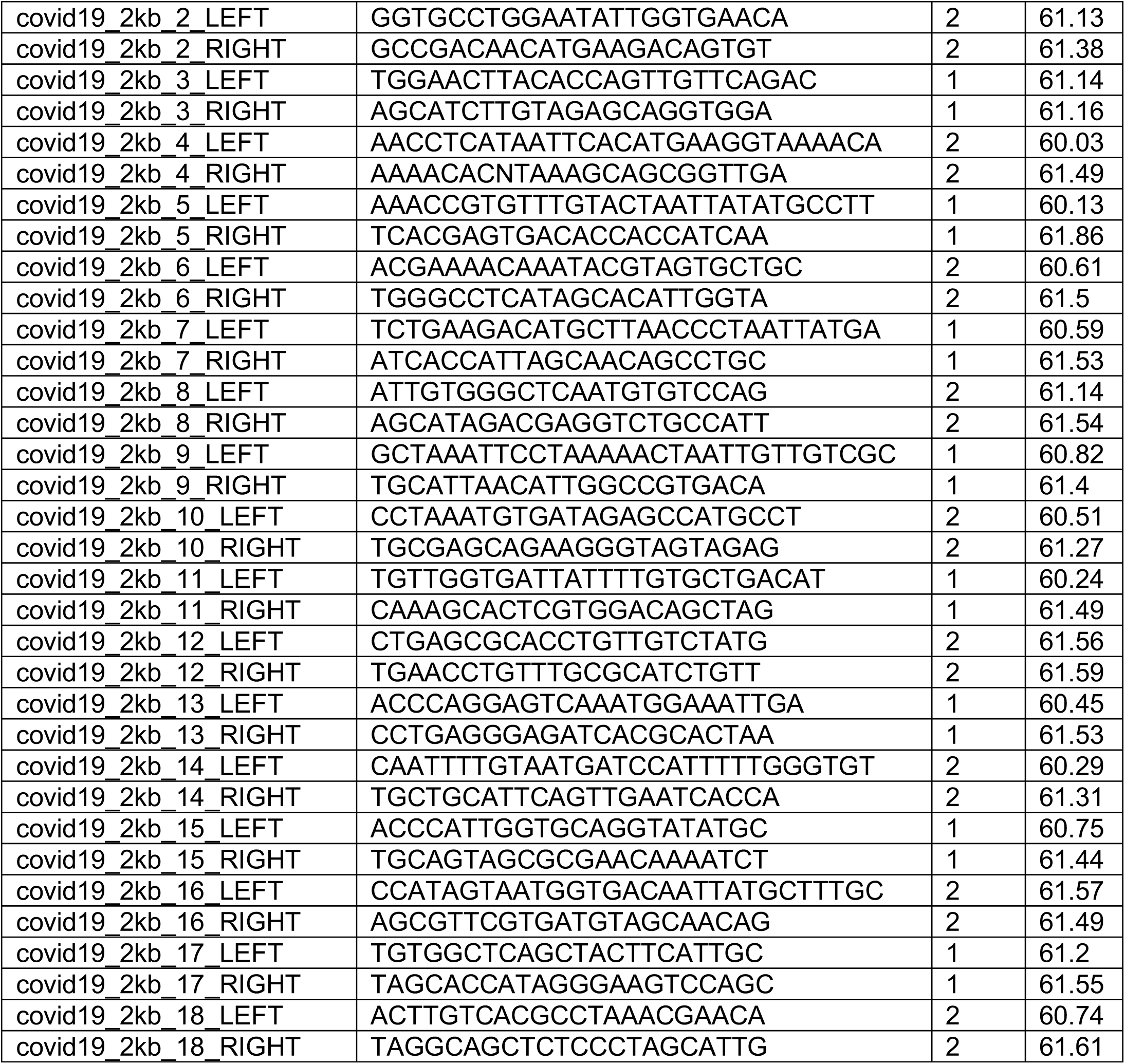
SARS-CoV-2 primer sets for whole-genome amplification

**Figure S1.**
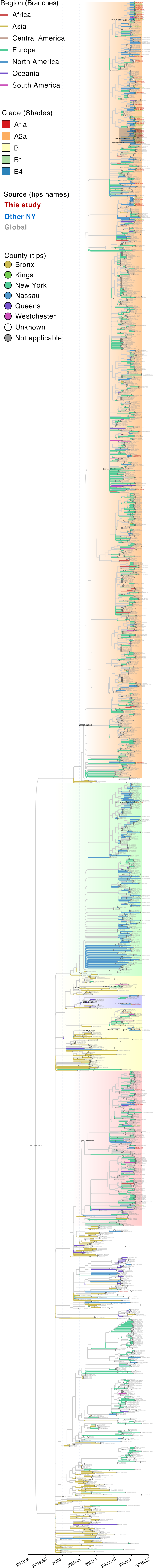
Detailed phylogenetic relationships of NY isolates (February 29 to March 18, 2020). Maximum likelihood time-scaled phylogenetic inference for 2,363 global SARS-CoV-2 sequences available in GISAID EpiCoV database as of April 1, 2020 and 84 NY sequences from this study. Branches are colored according to the region of origin, tip names are colored according to their source indicating the position of NY sequences in red (this study), blue (other NY sequences) and grey (other). Tip circles indicate NY state County assigned from zip code information (this study) and metadata from GISAID submissions. Clades that contain NY sequences and putative community spread clusters are highlighted with different colors where the grey shade on top of clade A2a indicates the position of the NY local transmission cluster (see main text for clade definition). The 90% confidence interval of the inferred time of the most recent common ancestor is indicated for the root and statistically supported nodes (>70% bootstrap values) related to sequenced isolates from NYC.

